# Early detection of SARS-CoV-2 infection cases or outbreaks at nursing homes by targeted wastewater tracking

**DOI:** 10.1101/2021.01.21.21249640

**Authors:** Laura Davó, Raimundo Seguí, Pilar Botija, María José Beltrán, Eliseo Albert, Ignacio Torres, Pablo Ángel López-Fernández, Rafael Ortí, Juan Francisco Maestre, Gloria Sánchez, David Navarro

**Affiliations:** Global Omnium, Valencia, Spain; Dirección de Atención Primaria, Departamento de Salud Clínico-Malvarrosa, Hospital Clínico Universitario de Valencia, Valencia, Spain; Dirección de Enfermería, Departamento de Salud Clínico-Malvarrosa, Hospital Clínico Universitario de Valencia, Valencia, Spain; Microbiology Service, Clinic University Hospital, INCLIVA Health Research Institute, Valencia, Spain; Department of Preventive Medicine and Quality Assurance, Clinic University Hospital, INCLIVA Health Research Institute, Valencia, Spain; Food Safety and Preservation Department, Institute of Agrochemistry and Food Technology (IATA-CSIC), Valencia, Spain; Department of Microbiology, School of Medicine, University of Valencia, Valencia, Spain

**Keywords:** SARS-CoV-2 RNA, Nursing homes, wastewater, near-source tracking, COVID-19 outbreak

## Abstract

**Objectives:** Near-source tracking of SARS-CoV-2 RNA in the sewage drains serving particular buildings may allow rapid identification of SARS-CoV-2 infection cases or local outbreaks. In this pilot study, we investigated whether this was the case for nursing homes (NH).

**Methods:** The study involved five NH (from A to E) affiliated to the Clínico-Malvarrosa Health Department, Valencia (Spain). These were nursing or mixed nursing/care homes of different sizes, altogether providing care for 472 residents attended by a staff of 309. Near-source sewage samples were screened for presence of SARS-CoV-2 RNA by RT-qPCR at least 5 days per week during the study period. SARS-CoV-2 RNA testing in nasopharyngeal swabs from residents and staff was performed with the TaqPath COVID-19 Combo Kit (Thermo Fisher Scientific, Massachusetts, USA).

**Results:** SARS-CoV-2 RNA was detected in wastewater samples from four of the five NH. SARS-CoV-2 infection cases were documented in three of these four NH. Of the two NH without SARS-CoV-2 infection cases, no SARS-CoV-2 RNA was detected in sewer samples from one facility, while it was repeatedly detected in samples from the other. Presence of SARS-CoV-2 RNA in sewage preceded identification of isolated cases among residents or staff or outbreak declaration in two NH, with lag times ranging from 5 to 19 days.

**Conclusion:** Our study demonstrated that intermittent or persistent detection of SARS-CoV-2 RNA in NH sewers can provide an early warning of subsequent individual cases or outbreaks in these facilities.

## INTRODUCTION

Nursing homes (NH) have been severely affected by the COVID-19 pandemic, largely due to their congregate nature and the vulnerability of residents [1,2]. Advanced age, frailty and concurrence of underlying chronic health conditions place NH residents at high risk for developing severe forms of COVID-19 and death. In Spain, at least 50% of NH-resident deaths officially reported by the Ministry of Health have been directly or indirectly attributed to COVID-19. Early detection of SARS-CoV-2 outbreaks at NH through periodic and systematic RT-PCR screening of residents and personnel has been invoked as a seemingly effective strategy to rapidly blunt virus spread in this setting [3,4]. Routine implementation of this approach has encountered many logistical obstacles, however, and to our knowledge its cost-effectiveness has not been incontrovertibly proven. Long-lasting virus shedding of SARS-CoV-2 in urine and feces has been documented in both symptomatic and asymptomatic infected adults [5]. As a result, near-source tracking in the sewers serving particular buildings has emerged as an appealing non-invasive tool which when combined with subsequent targeted population screening when SARS-CoV-2 is detected may enable rapid identification and control of facility outbreaks [6,7].

In this pilot study, we provide evidence demonstrating the feasibility and utility of this wastewater-based epidemiological approach for early identification of isolated cases or outbreaks of SARS-CoV-2 infection in NH.

## MATERIAL AND METHODS

### Nursing homes and sewage sampling

This pilot study involved five NH (listed as A to E) facilities located in Northeast Valencia (Spain), affiliated to the Clínico-Malvarrosa Health Department. These were nursing or mixed nursing/care homes of different sizes (Table 1), altogether providing care for 472 residents attended by 309 staff. Selection from among the 17 NH supported by the Clínico-Malvarrosa Health Department was based upon two criteria: (i) existence of sewage drain(s) not shared with nearby buildings and (ii) personal autonomy of most residents. NH sewage drain(s) were monitored for presence of SARS-CoV-2 RNA by testing near-source wastewater samples at least 5 days per week from October 7 to December 28, 2020. All except one NH (NHE) had a single drain site. Grab samples were collected on site from water outlets at each facility. At NHE, hierarchical pooling involved testing a combination of multiple samples from different sampling sites. Positive pool samples were deconvoluted and individually tested. All samples were taken early in the morning, collecting 1 L of water in sterile plastic containers with sodium thiosulfate (VWR, USA). Water samples were transferred to the laboratory, refrigerated at 4 °C and concentrated within 24 h.

**Table 1.**
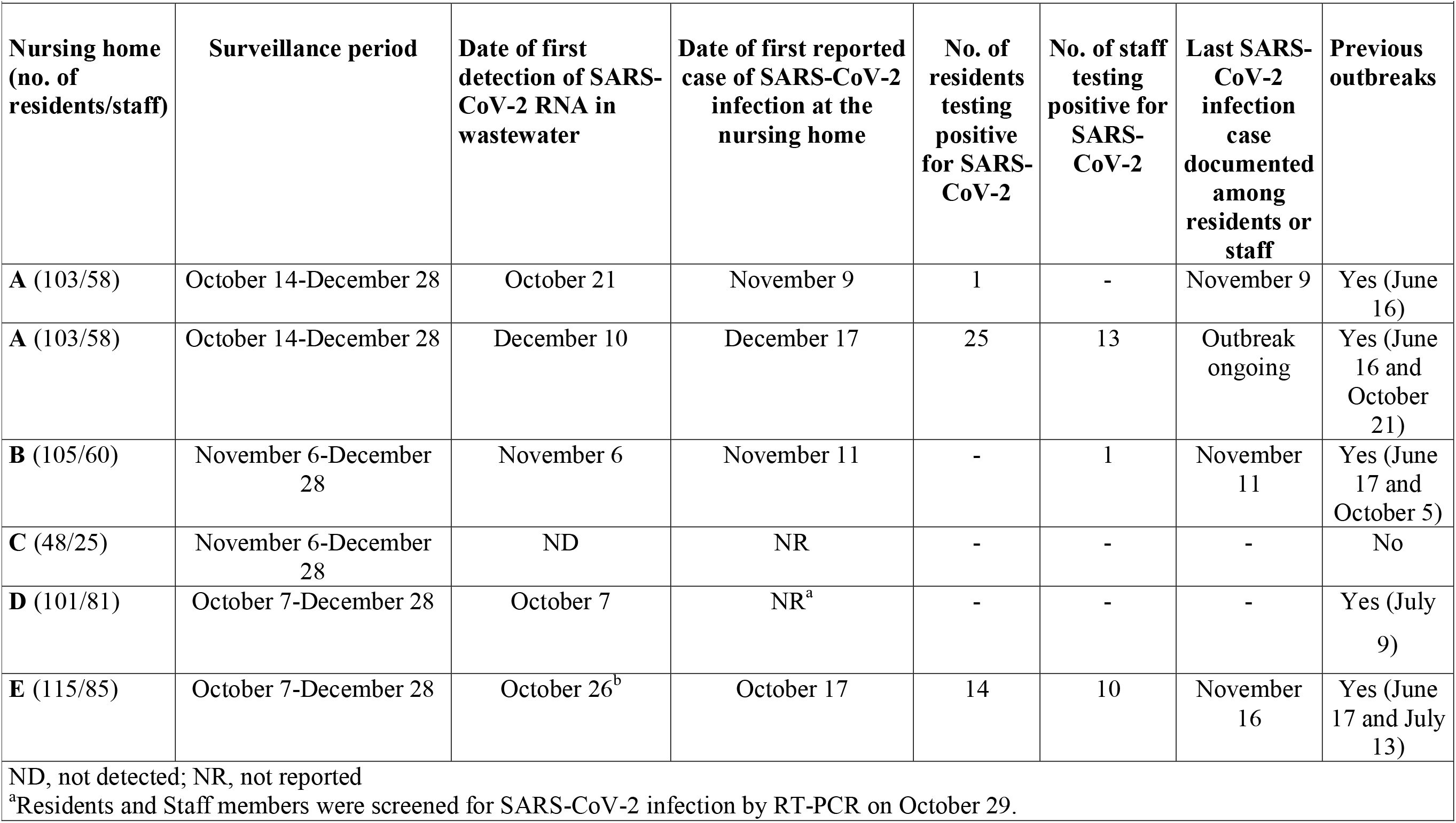
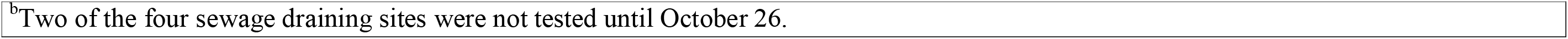
**Detection of SARS-CoV-2 in wastewater, residents and staff at nursing homes included in the study**

| **Nursing home**<br>**(no. of**<br>**residents/staff)** | **Surveillance period** | **Date of first**<br>**detection of SARS-**<br>**CoV-2 RNA in**<br>**wastewater** | **Date of first reported**<br>**case of SARS-CoV-2**<br>**infection at the**<br>**nursing home** | **No. of**<br>**residents**<br>**testing**<br>**positive**<br>**for SARS-**<br>**CoV-2** | **No. of staff**<br>**testing**<br>**positive for**<br>**SARS-**<br>**CoV-2** | **Last SARS-**<br>**CoV-2**<br>**infection**<br>**case**<br>**documented**<br>**among**<br>**residents or**<br>**staff** | **Previous**<br>**outbreaks** |
| --- | --- | --- | --- | --- | --- | --- | --- |
| **A** (103/58) | October 14-December 28 | October 21 | November 9 | 1 | - | November 9 | Yes (June 16) |
| **A** (103/58) | October 14-December 28 | December 10 | December 17 | 25 | 13 | Outbreak ongoing | Yes (June 16 and October 21) |
| **B** (105/60) | November 6-December 28 | November 6 | November 11 | - | 1 | November 11 | Yes (June 17 and October 5) |
| **C** (48/25) | November 6-December 28 | ND | NR | - | - | - | No |
| **D** (101/81) | October 7-December 28 | October 7 | NR<sup>a</sup> | - | - | - | Yes (July 9) |
| **E** (115/85) | October 7-December 28 | October 26<sup>b</sup> | October 17 | 14 | 10 | November 16 | Yes (June 17 and July 13) |

ND, not detected; NR, not reported
<sup>a</sup>Residents and Staff members were screened for SARS-CoV-2 infection by RT-PCR on October 29.
<sup>b</sup>Two of the four sewage draining sites were not tested until October 26.

### SARS-CoV-2 detection and quantitation in sewage samples

Sewage water samples were analyzed at Global Omnium laboratory. Samples were concentrated using the aluminum adsorption-precipitation method [8,9]. A final concentrate was then obtained by centrifugation at 1,900 × g for 30 min; the resulting pellet was resuspended in 1 mL of PBS, pH 7.4. Viral extraction from wastewater concentrates was carried out using the NucleoSpin RNA virus Kit (MACHEREY-NAGEL, Germany). SARS-CoV-2 RNA detection was performed by RT-qPCR using One Step PrimeScript™ RT-PCR Kit (Perfect Real Time) (Takara Bio, USA), targeting the nucleoprotein (N), N1 and N2 fragments [10], and envelope protein (E) gene [11]. RNA samples were analyzed in duplicate. Each RT-qPCR run included negative (nuclease-free water) and positive controls. RT-qPCR targets were quantified by plotting the quantification cycles (C_T_) to an external standard curve built with 10-fold serial dilution of the 2019-nCoV_N_Positive Control and 2019-nCoV_E_Positive Control (IDT). Mengovirus RNA recovery rates were calculated and used as quality assurance parameters according to ISO 15216-1:2017 [12]. Results are reported as genome copies (GC)/L.

### SARS-CoV-2 testing in residents and staff

Nasopharyngeal swabs (NP) for RT-PCR testing were collected by experienced nurses at the NH sites and immediately placed in 3 ml of Universal Transport Medium (UTM, Becton Dickinson, Sparks, MD, USA). RT-qPCRs were conducted within 24 h of specimen collection at the Microbiology Service of Hospital Clínico Universitario (Valencia, Spain) with the TaqPath COVID-19 Combo Kit (Thermo Fisher Scientific, Massachusetts, USA) [13]. RNA was extracted using the Applied Biosystems™ MagMAX™ Viral/Pathogen II Nucleic Acid Isolation Kits coupled with Thermo Scientific™ KingFisher Flex automated instrument (Thermo Fisher Scientific).

### Ethics statement

Permission to analyze the wastewater was granted by the nursing home operator and local authority responsible for the sewer system. Ethical approval for this study was waived by the Hospital Clínico Universitario INCLIVA Ethics Committee because RT-PCR testing either for diagnosis purposes or surveillance of both nursing home residents and staff are usual practices at health Department Clínico-Malvarrosa, Valencia, Spain.

## RESULTS

SARS-CoV-2 RNA was detected in wastewater samples collected from four out of the five NH. The dynamics of SARS-CoV-2 RNA detection and viral loads measured at each NH are shown in Figure 1. SARS-CoV-2 infection cases, either asymptomatic or symptomatic, were documented in three of the four NH (Table 1). No cases were identified at NHD within the study period, despite repeated detection of SARS-CoV-2 RNA in the sewage drain. Of note, residents and staff at NHD were screened by RT-PCR on October 29, twelve days after SARS-CoV-2 was first detected in sewage, all yielding negative results. SARS-CoV-2 RNA was not detected in samples collected from NHC and no cases were documented throughout the follow-up period. The timespan between first SARS-CoV-2 RNA detection in sewage and index case diagnosis at each NH is shown in Table 1 (and depicted in Figure 1). Presence of SARS-CoV-2 RNA in sewage preceded identification of isolated cases among residents or staff (in both cases symptomatic) or outbreak declaration in two NH (NHA at during two different time periods, and NHB). Repeated detection of SARS-CoV-2 RNA was not documented until after outbreak declaration in the case of NHE (Table 1), although it should be noted that between October 7 and first case detection on October 17 only two of the four sewage drains at NHE had been sampled.

**Figure 1.**
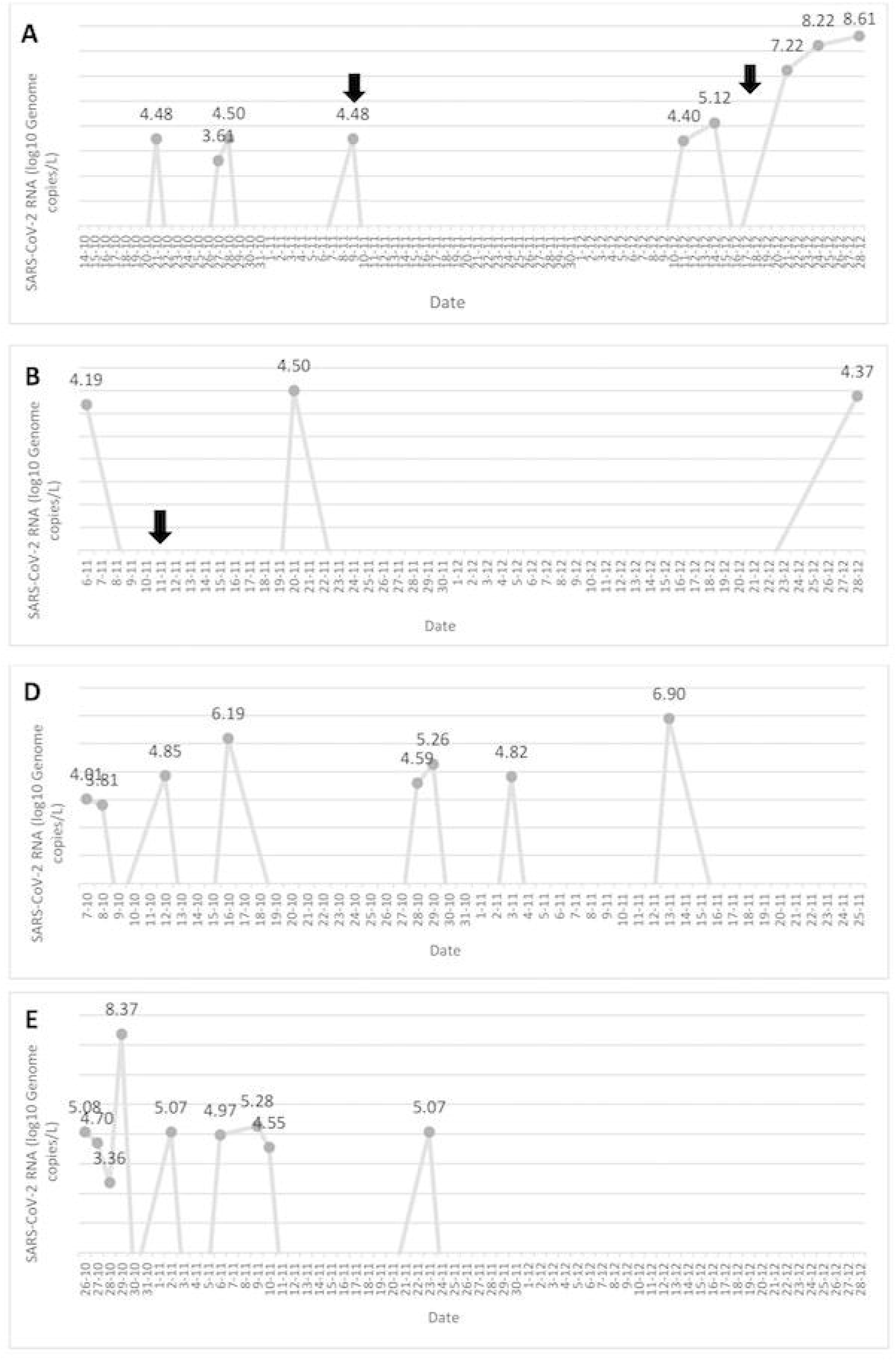
Dynamics of SARS-CoV-2 RNA detection in sewage drains of nursing homes (NH). SARS-CoV-2 RNA loads (log_10_ genome copies-GC-)/L) at each time point are mean values of duplicate measurements at NH with a single sewage drain (NHA, NHB, NHC, NHD) or mean values quantified at the different sampling sites of the facility (NHE). Arrows point to the date of first detection of cases among residents or staff at a given NH.

SARS-CoV-2 RNA levels in wastewater samples increased exponentially over the course of NH outbreaks (NHA and NHE), reaching peak levels above 8.0 log_10_ GC/L (Figure 1).

Finally, disappearance of SARS-CoV-2 RNA from sewers was associated with control of outbreaks or absence of new case documentation following implementation of adequate measures (isolation of positive case and quarantining of close contacts) (Figure 1). The SARS-CoV-2 outbreak at NHA is currently still active.

## DISCUSSION

Wastewater SARS-CoV-2 RT-PCR testing has emerged as an efficient strategy for epidemiological surveillance of COVID-19, as traces of SARS-CoV-2 RNA frequently predate detection of cases in the community by between 4 and 15 days [8,14-16]. Likewise, tracking SARS-CoV-2 in sewer systems from different facilities (i.e. campus dorms, workplaces, correctional facilities, schools) may allow early documentation of SARS-CoV-2 circulation, thus potentially contributing to prompt blunting of viral transmission [6,7]. Results from this study suggested that SARS-CoV-2 RNA surveillance of sewage drains may indeed serve as an early warning system for isolated cases or outbreak declaration of SARS-CoV-2 infection in NH. This was found to be the case in NHA and NHB with lag times ranging from 5 to 19 days, and although speculative, could also have been the case in NHE, had we not missed two out of the four sewage drains at NH. Of analogous importance was the fact that SARS-CoV-2 RNA traces were not detected in NHC, the facility in which no cases were reported within the study period.

An intriguing observation was that repeated detection (at 8 time points) of SARS-CoV-2 in NHD sewers was neither preceded nor followed by case detection in this facility. Furthermore, all residents and staff at NHD were screened by RT-PCR as part of this pilot experiment a few days after first SARS-CoV-2 RNA detection in sewage, all returning negative results. Although a plausible explanation for this was that precautionary measures to avoid virus transmission were maximized following first detection of SARS-CoV-2 RNA in this particular facility, and that any infected resident/s or staff member/s may have yielded false-negative RT-PCR results [17], a thorough investigation conducted after systematic screening of NHD population revealed the existence of cross-contamination between sewage drains of this NH and that of an adjacent building, that had gone unnoticed.

On the other hand, as expected, SARS-CoV-2 RNA levels in sewage drains increased dramatically during outbreak periods.

The main limitation of the current study is the relatively limited number of NH recruited for the study.

In conclusion, this pilot study proved that intermittent or persistent detection of SARS-CoV-2 RNA in NH sewage drains often anticipates declaration of individual cases or outbreaks. Frequent SARS-CoV-2 RT-qPCR sewage testing coupled with targeted screening of residents and staff may prove useful for early blunting of virus transmission and spread at NH. Further studies with a larger site sample are warranted to confirm this assumption.

## Data Availability

The data that support the findings of this study are available on request from the corresponding author

## ACKNOWLEDGMENTS

We thank all personnel working in the nursing homes and the Hospital Clínico Universitario of Valencia for their unwavering commitment in the fight against COVID-19.

## AUTHOR CONTRIBUTIONS

LD, RS, EA, IT: methodology and data validation. LA, RS, JFM, GS and DN: formal analysis. LD, RS, JFM, GS and DN: Conceptualization, supervision. PB, MJB, PL-F and RO: supervision of RT-PCR testing at NH facilities. LD, GS and DN: writing the original draft. All authors reviewed and approved the original draft.

